# Climate change, national vulnerability and personal anxiety among teenagers and adults in 108 countries: an instrumental variable analysis

**DOI:** 10.1101/2025.05.01.25326816

**Authors:** Gindo Tampubolon

## Abstract

The latest Assessment Report from the Intergovernmental Panel on Climate Change (2022) stated for the first time “with high confidence [that] climate change is human made.” However, this scientific fact has not elicited widespread belief even among democratic representatives, let alone its mental health impact being universally recognised.

Using the latest survey in 2020 of extreme anxiety among 113,460 teenagers and adults in 108 countries which constitute 89% of the world population, I ask whether personal belief in climate change drives extreme anxiety. Such a causal link arises when urgent actions endorsed by the Panel are only slowly heeded by governments while personal lifestyle changes are perceived as hardly sufficient, creating chronic dissonance.

But there may be other drivers not elicited in the survey eg, personal action on climate change and exposure to misinformation (eg: a democratic representative claims that government geoengineers hurricanes which harm health, instead of their increased frequency being a manifestation of climate change). To overcome potential bias, I use fixed effect instrumental variable analysis (instruments: country average belief in science today and two years previously). This is complemented with a placebo test, heterogeneity analysis across gender, sensitivity analysis (random effect) and mechanistic analysis across nations with different levels of vulnerability.

I found that personal belief in climate change raises the probability of extreme anxiety by 12% (confidence interval 6–17%). And national context matters a great deal. Together they suggest that the less-than-urgent actions on climate by governments already harm mental health around the world. The nexus of effective climate action by governments and health benefits for citizens represents a synergy between two sustainable development goals which commend themselves to immediate action.

## INTRODUCTION

The Intergovernmental Panel on Climate Change made a strong shift in its Sixth Assessment Report (AR6) by stating “with high confidence that climate change is human made”.^1^ This conclusion marked a crucial point in the scientific consensus on climate change, attributing the majority of observed global warming since the mid-20th century directly to human activities, including the burning of fossil fuels and deforestation. While earlier reports by the panel of leading scientists suggested that human influence was “likely” responsible for climate change, AR6 left no room for ambiguity, calling for urgent actions and large-scale collective interventions.

Climate change is a quintessential scientific fact, arrived at through systematic observations and open refutation by scientists around the world.^2–4^ It need not tally with personal experience at any one time. Our personal quotidian experience can so easily lead to doubt of the fact of climate change which includes global warming, an upward secular trend in world temperature unlike that known before human activities predominate from the Industrial Revolution onwards. One can experience a cold spell on a summer’s day with non-negligible probability. Scientists establish the claim that even with this non-negligible probability, the world is indeed climbing a higher temperature trend. For example, here is the famous Keeling curve I fetched while writing this manuscript, an important continuous measurement of carbon dioxide concentration in the atmosphere taken in the same place at Mauna Loa for nearly 70 years, which helps to establish the scientific consensus on climate change. It is in principle and practice open to access by anyone with a smartphone, including scientists and the public, hence open to refutation by the same. The curve shows the complex, systematic and patterned increase in carbon dioxide concentration in the atmosphere. For the last 70 years or so there is a secular increase in this concentration (I superimpose a line fit), and each year the Earth exhibits a breathing pattern with a one year cycle and peak exhalation in May.

The fact of climate change is increasingly akin to the scientific facts of smoking tobacco being a major health risk to anyone or of driving under the influence of alcohol being a major risk to road users. It need not tally with our personal experience, perhaps because having believed in it, we now avoid smoking or drinking-and-driving. The quintessentially scientific and complex nature of climate change means that personal beliefs in science and in climate change matter. This work thus explores the effect of such personal belief in climate change on personal mental health as one of the impacts.

AR6 emphasizes that these impacts — manifesting in heatwaves through flash floods to other extreme weather events — are becoming more severe and more frequent. It also stresses that these effects are already being felt across all regions of the world, with vulnerable communities bearing the brunt. The science is unequivocal: without immediate and coordinated global actions, the consequences of climate change could become irreversible. The Report goes beyond the science of climate change, putting a stark warning about the narrowing window of opportunity for meaningful action to mitigate its worst effects. The Panel’s urgent demand is for collective global action — specifically from governments, businesses, international institutions and persons— to dramatically reduce greenhouse gas emissions and transition to sustainable systems. AR6 highlights that personal actions,^5^ while important, are insufficient to curb the scale of the crisis. It calls for systemic change, emphasizing policy reforms, large-scale infrastructure overhauls, and global cooperation. For instance, transitioning energy systems away from fossil fuels toward renewable energy, rethinking land use and agriculture, and inventing green technologies are framed as non-negotiable steps. If these actions are merely slowly heeded, the Earth’s capacity to remain hospitable to human life will be severely compromised for our common future.^6^

Yet, despite these warnings and clear demands for global-scale solutions, the political and economic responses to AR6 have been notably inadequate.^7^ While some progress has been made, such as international agreements like the Paris Climate Accord,^7^ the implementation of transformative policies remains slow, fragmented, and often driven by short-term interests rather than long-term sustainability. Major polluting nations and corporations have resisted making the necessary sacrifices or investments, citing economic concerns, geopolitical competition, or energy security. The gap between the scale of action needed and the actions taken is widening, further exacerbating the climate crisis and creating possible chronic dissonance in people.

This continuing slow action at a systemic level contrasts with the rising concern at the individual level. More and more people are aware of the tangible effects of climate change and feel an increasing sense of personal responsibility.^5^ However, the Panel’s clear message that personal actions — such as reducing one’s carbon footprint or switching to eco-friendly products — are insufficient to mitigate climate change can lead to feelings of powerlessness. The disconnect between personal responsibility and the systemic action required can lead to anxiety.

This discussion raises an important question: can personal belief in the reality and risks of climate change be linked to heightened feelings of anxiety? While environmental worry can be seen as a rational response to the crisis, the burden of feeling responsible, combined with the helplessness of witnessing insufficient global action, may create significant psychological stress. This hypothesis posits that individuals who believe in climate change and its impacts may be at a higher risk of experiencing extreme anxiety, particularly as they confront the widening gap between personal efforts and the lack of required collective action.

The scientific fact of climate change is systematic and replicable like the Keeling curve while personal belief is contingent. The mechanism which gives rise to the outcome of extreme anxiety is contingent on government or more broad collective actions. The Panel has recommended a raft of urgent actions, and when the actions are delayed, those who believe in the risks of climate change may experience dissonance and anxiety. This can manifest in general anxiety i.e. not only when primed or phrased with climate change issues in the news or in conversations.

AR6 and other works detail how climate change manifestations including heatwaves and flash floods are associated directly and indirectly with physical and mental health.^8^ These major risks to health are now well recognised and recent works have moved towards capturing a specific psychological construct of anxiety related to facing this global crisis, for instance by devising the climate change anxiety scale.^9–11^ Using the scale, a study has found that climate anxiety is predicted by personal attributes, showing higher prevalence among young Britons.^11^ Recent studies of climate anxiety using this or other scales recognise that climate anxiety can be either a mental health condition or a rational adaptation to the known global crisis. These studies often focus on the developed countries due to the greater media attention, research, and resources dedicated to health in those regions. However, this does not mean that climate anxiety is absent in developing countries. In fact, the experience of climate change in developing nations can be even more immediate and devastating, given the heightened vulnerability of their populations to climate impacts as amply documented in AR6 and elsewhere.^1,12^ The mental health impacts in these regions may be shaped differently, often intertwined with daily survival, economic instability, and direct exposure to global warming-related disasters and diseases. For example, the World Health Organization conference on Health and Climate since 2014 pointed out that developing countries are highly vulnerable to the health effects of climate change, including mental health impacts.^12^ Further reports emphasize that people in low-income countries may experience higher levels of anxiety and stress due to the increased frequency of climate-related disasters, food insecurity, and displacement (ibid). The Lancet Countdown on health and climate change in its 2024 report highlighted how the climate crisis disproportionately affects low-income countries, contributing to mental health issues (lancetcountdown.com accessed 21 Apr 2025).

But my focus here is on general i.e. extreme anxiety which is elicited with one item without being primed by climate change issues.^13^ This is obtained by a single affirmative question on anxiety, eliciting a yes or no response. A new aspect here is the search or the hypothesis for its cause in the belief in climate change. *A note on terminology*. Although climate change is broader than global warming, the two are presented as equivalent to the participants in 108 countries in this way, “do you believe that climate change or global warming poses major risks?” eliciting a yes or no response. *Pace*, climate change includes sea level rises and rainfall pattern changes (<u>NASA</u>. Accessed Dec 2020.) Further, because AR6 has high confidence in the scientific claims that climate change is human made and that it poses myriad risks, belief in climate change or global warming is used interchangeably.^1,2^

The mechanism connecting belief in the scientific fact and extreme anxiety arises from the dissonance of seeing inadequate responses (of national, international, business and individual actions) to shore up our resilience and reduce our vulnerability by deploying required adaptation and mitigation actions. Without commensurate collective actions, in turn conditioned by regulation and incentives, the belief in climate change posing major risks can become mentally deleterious.

To preview two key results: first, the data strongly suggest that personal belief in climate change raises the probability of experiencing extreme anxiety, sufficient to hinder normal day to day activities for a fortnight. Second, the posited mechanism also receives support from the data when augmented with the Panel’s own independent assessment of national vulnerability.^1^ When effective urgent actions are taken by some nations to make them less vulnerable or more resilient, their citizens’ personal belief in climate change raises much less extreme anxiety. Note that taking the first result as implying “to preserve mental health one should doubt climate change” amounts to a counsel of despair. Instead, the second key result implies that nations should intensify effective collective actions to adapt and mitigate against climate change if we want to make nations resilient and help citizens to reduce extreme anxiety.

This work makes a number of contributions to the literature on climate change, global health and global development. Personal beliefs in science in general and in climate change in particular are already known as important constructs in public policy and public life today.^3,9,14–16^ These results add to and enrich this venerable literature with new evidence on the link between personal belief in climate change and mental health. The results also speak to the interlink between two sustainable development goals (urgent actions on climate change and health for all) specifically showing that the two are synergistic rather than contradictory.^17^ This is a critical demonstration of worldwide scope.

## MATERIALS

### Wellcome Trust Global Monitor Outcome and Treatment

The data were drawn from the latest Wellcome Global Monitor which surveyed 113,460 teenagers and adults aged 15 to 99 across 108 countries or 89% of the world population in 2020.^18^ The outcome is personal anxiety, assessed through a single binary question developed by an international team of experts, including those from the World Health Organization, Harvard University, London School of Hygiene and Tropical Medicine, London School of Economics and Political Science and the Wellcome Trust. The survey defines “extreme anxiety or depression” as a condition where a person feels so anxious or depressed that she or he is unable to continue regular daily activities for two weeks or longer. Although the item has not been extensively validated, it nevertheless was devised with extensive expert inputs which consider this quality against the feasibility of use in more than 100 countries.^18^ Participants were also asked to indicate their belief in climate change by responding to whether they believe “climate change or global warming is a major risk,” eliciting a yes or no response. Belief in climate change serves as the treatment variable.

While anxiety may be influenced by belief in climate change, it is also determined by other factors. Important personal factors include sex (male and female), age, income (in quintiles), employment status (unemployed, part-time, full-time), education level (primary, secondary, or tertiary) and use of social media (0: never to 6: several times an hour) – common factors in empirical studies of mental health and wellbeing.^19–23^

### Analytic strategy

To test the hypothesis that personal belief in climate change raises the risk of extreme anxiety, I use a fixed effect instrumental variable model in Stata (command xtivreg2) because it gives robust estimates of the causal effect of personal belief in the presence of omitted variables and it does not assume that country invariant attributes are uncorrelated with the error term.^24^ If this last assumption were made, then a random effect instrumental variable model could be applied which in turn admits context or country factors (more on this below in the sensitivity analysis). Meanwhile, other benefits of the chosen model include an extensive battery of tests: endogeneity test, relevance test and overidentifying restriction test.^25–28^ All these outweigh the perceived disadvantage of applying the model to explain a binary dependent variable. Unfortunately, the anxiety duration in number of days was not elicited which would have accorded with the model or command. Moreover, there is no analogous fixed effect instrumental variable command for binary dependent variables. As an upside, the chosen model gives coefficients that can be interpreted as marginal probabilities.

The instruments are country average of belief in science two years previously (low or high, split at the median) and analogously in the current year. The Wellcome Global Monitor measured responses to the prompt of belief in science as “the understanding we have about the world from observation and testing, by people who study Planet Earth, nature, medicine, among other fields.”^13^ The previous sample was an independent sample so all the participants were included in constructing the average; the current year construction excludes the focus participant. For example, for participant *i* in country *j* the current year average is constructed by excluding participant *i* while including all other participants in the country.

Extensive complementary analyses are also conducted including random effect instrumental variable model (command xtivreg),^24^ stratified analyses based on sex, placebo test and mechanism analysis. The placebo test can be explained in reference to the directed acyclic graph below (omitting covariates to achieve concision) which shows an instrumental variable model where Z: instrument (country average of belief in science), T: treatment (belief in climate change) and Y: outcome (extreme anxiety).

The exclusion assumption of the instrument is expressed by the absence of a diagonal arrow from the instrument to the outcome, essentially an untestable assumption.^25–28^ However, its plausibility can be shown by a placebo test which here proceeds by replacing the treatment (belief) with a placebo viz. the belief of another random person drawn from the same country. The placebo must have no effect.

### Context in sensitivity analysis

As part of the sensitivity analysis, a random effect model was fitted which admits context or country factors. The survey’s global reach provides a context for analysing the association between country factors and anxiety. Key among them is the North-South distinction. Countries in the Global South are at greater risk of climate impacts for three main reasons. They have fewer resources to fund adaptation and mitigation efforts. They often depend more on natural services (e.g. agriculture) which employ a larger portion of the population and are more susceptible to climate impacts. They are home to families with less financial capacity for personal adaptation and mitigation actions. To account for this context, both North-South distinction and average income (per person) are included.

Importantly, during the initial administration of the survey in early 2020 it quickly became clear that the unfolding covid-19 pandemic would force radical change to the survey operation. Eventually, it turned into phone interview mode without face to face elicitation.^18^ This has specific relevance for an investigation explaining personal anxiety such as this study because the first half of 2020 was a period when the world began to come to terms with the unfolding pandemic. This was a shock to general mental health around the world. To capture this shock I included for each country the total number of cases during the period per million population (log), reasoning that higher number of cases would correlate positively with increased anxiety (Our World in Data, 2024, accessed 1 Nov 2024).

Mechanism analysis is implemented by expressing the logic which gives rise to personal anxiety. Recall that the hypothesis is motivated by the gap between personal belief in climate change and inadequate collective actions by governments, businesses and other institutions. I use an index of national vulnerability (low or high, split at the median), where *vulnerable* nations (high vulnerability) are those that have not yet managed to effectively prepare themselves to tackle the risks of climate change according to the IPCC. The index was developed in AR6 Working Group II which captures exposure, susceptibility, coping and adaptation capacity, altogether summarising more than just structural but also social and economic dimensions. The data and documentation are drawn from the IPCC (https://ipcc-browser.ipcc-data.org/browser/dataset?id=3736, accessed 1 Nov 2024). For concise interpretation, low vulnerability nations are deemed *resilient*. The mechanism suggests that in vulnerable nations where the actions were inadequate the effect of personal belief will be accentuated and strong, whereas in resilient nations where effective actions were in place the effect will be significantly weakened. To implement this mechanism an interaction term between national vulnerability and the treatment is included.

## RESULTS

I begin by showing a map drawing the average level of extreme anxiety reported across different countries around the world (figure 3). The colour-coding provides a visual representation of the geographical patterns. The majority of countries in North America, Europe, and parts of East Asia are shown in lighter shades, indicating relatively lower average levels of anxiety and depression. In contrast, many parts of Africa, Latin America, and Asia exhibit much darker hues, suggesting higher reported rates of these mental health problems.

**Figure 1.**
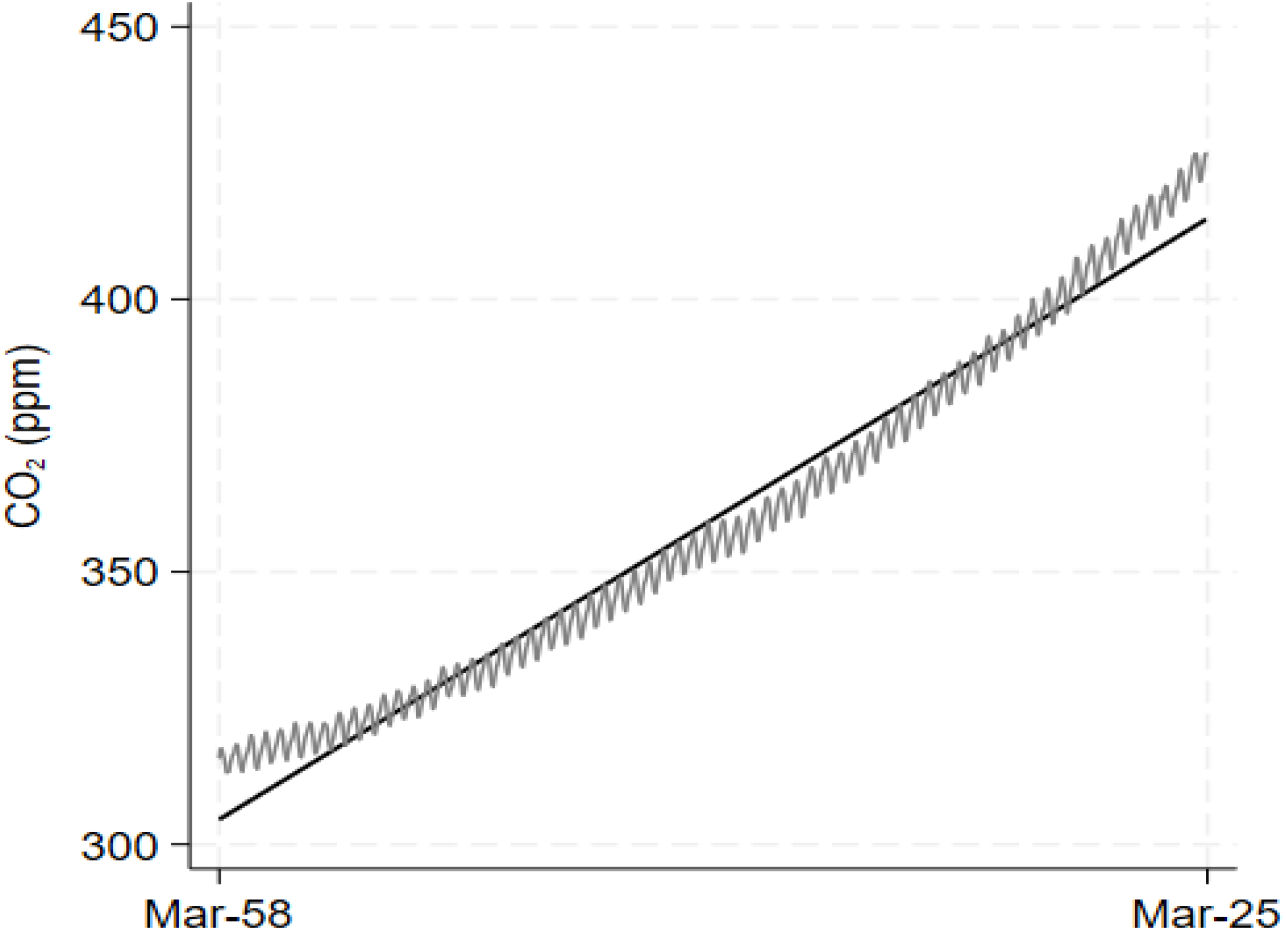
The Keeling curve of carbon dioxide concentrations (monthly, parts per million) in the atmosphere (1958 to 2025). Source: Scripps Institution of Oceanography, Author.

**Figure 2.**
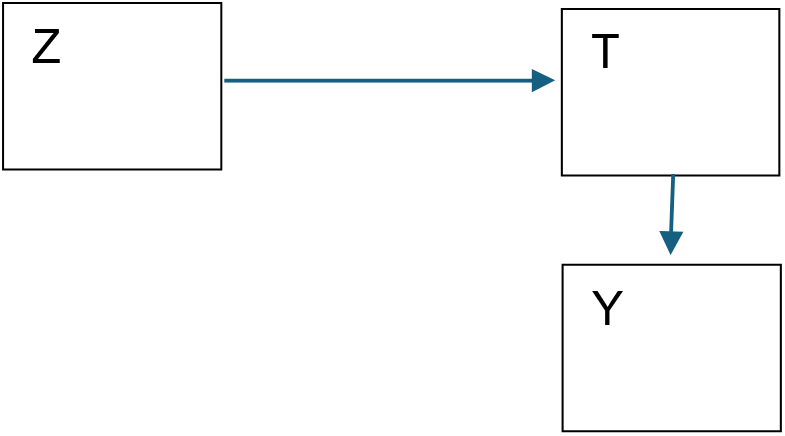
Directed acyclic graph of an instrumental variable analysis

**Figure 3.**
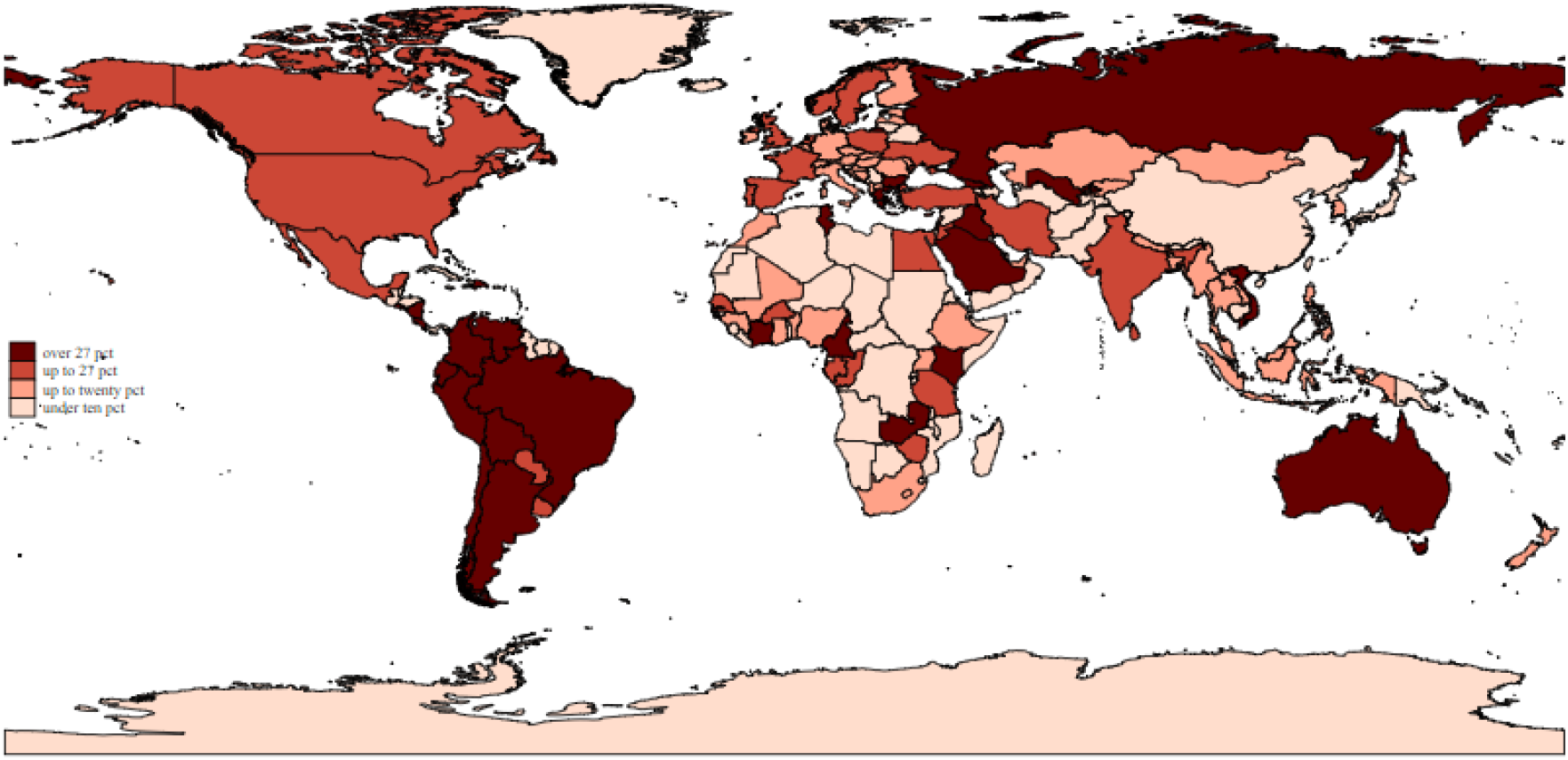
Map of extreme anxiety around the world in 2020. Source: Wellcome Global Monitor (2023).

Drilling deeper, we can see pockets of high anxiety/depression even within the generally lower-scoring regions of the Global North. For instance, certain parts of Southern Europe, such as Greece and Spain, appear to have higher average levels compared to Northern European countries. It is important to note that this map represents country average data and may mask important individual variations. Additionally, differences in reporting and cultural perceptions of mental health could influence the observed patterns to some degree, although the Wellcome Report question has undergone some testing to minimize this influence. Potential drivers of these global and regional variations may include socioeconomic factors, access to mental health resources and exposure to environmental or geopolitical stressors. For example, the high-anxiety regions in parts of Africa and Latin America could be linked to political instability, territorial conflict, or economic challenges that contribute to increased mental health burdens. This world map conveys a compelling visual representation of the global landscape of anxiety, highlighting the significant disparities that exist between countries.

I then explore the associations between this outcome of extreme anxiety and belief in climate change, plotting country averages and bringing in the distinction between them (vulnerable vs resilient) which is posited to be important in the mechanism. See figure 4.

**Figure 4.**
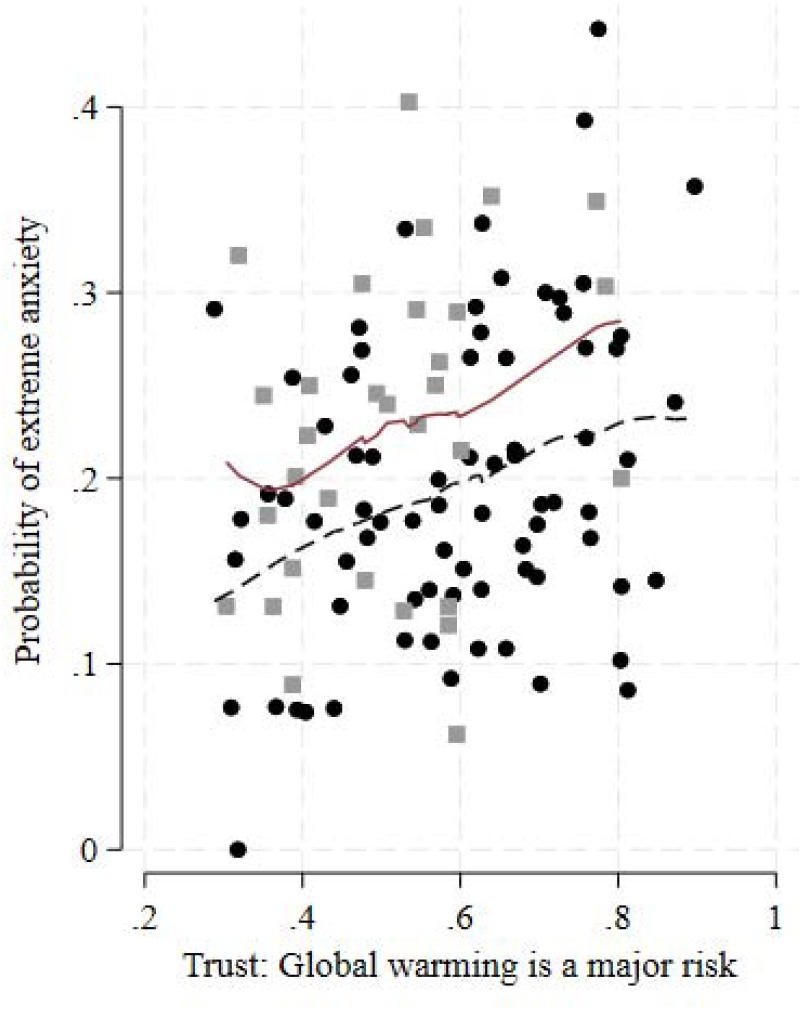
Extreme anxiety by belief in climate change (country averages), separating vulnerable (dots) from resilient (squares) countries. Source: Wellcome Global Monitor (2023).

The solid dots and solid line represent highly vulnerable nations. The plot shows a clear positive association between belief in climate change and anxiety. As belief in climate change increases, the probability of experiencing extreme anxiety also rises markedly. This indicates that in nations more susceptible to climate impacts, people who trust in the severity of global warming are more likely to experience higher levels of anxiety. Next, the squares and dashed line represent resilient nations. Similarly, there is a positive association between belief in climate change and anxiety, though the increase in anxiety with trust is less steep compared to that in vulnerable nations. This suggests that even in resilient nations, higher trust in the risks of global warming is associated with increased anxiety, but the effect is somewhat moderated compared to that among vulnerable nations. As summaries, the solid and dashed lines provide a smoothed fit to the data, reinforcing the observed trends. The solid line for vulnerable nations shows a steeper slope, indicating a stronger relationship between belief in climate change and anxiety. The dashed line for resilient nations, while still positive, is less steep, suggesting a more moderate relationship. The combined plot highlights that belief in the risks of global warming is positively associated in all nations. However, the impact is higher and more pronounced in vulnerable nations. Compared to the dashed line, the solid line is shifted upward and tilted upward, too. For the same level of belief in climate change, proportionately more people in vulnerable countries report extreme anxiety. Moreover, as the level of belief increases, for the same amount of increase in belief, proportionately even more people in vulnerable countries report extreme anxiety.

Already, this plot indicates that personal attributes do not explain personal anxiety around the world in a straightforward manner. To explain personal anxiety better, I use a fixed effect instrumental variable model on the Wellcome Global Monitor sample whose descriptive summaries are given in table 1. The coefficient estimates are collected in table 2, where three models are put together, for fixed effects model and random effects model as sensitivity analysis (penultimate column) as well as a placebo test model (last column).

**Table 1.** Descriptive summaries of the analytic sample – number (percentages) or mean (standard deviation). Source: Wellcome Global Monitor (2023).

|  | *Male*<br>N=57967 | *Female*<br>N=55493 | *Total*<br>N=113460 |
| --- | --- | --- | --- |
| *Been anxious or depressed* |  |  |  |
| No | 47053<br>(81.2%) | 43333<br>(78.1%) | 90386<br>(79.7%) |
| Anxious | 10914<br>(18.8%) | 12160<br>(21.9%) | 23074<br>(20.3%) |
| *Global warming is major risk* |  |  |  |
| Denier | 25647<br>(44.2%) | 23800<br>(42.9%) | 49447<br>(43.6%) |
| Believer | 32320<br>(55.8%) | 31693<br>(57.1%) | 64013<br>(56.4%) |
| *Age* | 39.1 (16.3) | 40.3 (17.2) | 39.7 (16.8) |
| *Education Level* |  |  |  |
| Elementary or less ($\leq 8$ years) | 7889<br>(13.6%) | 7852<br>(14.1%) | 15741<br>(13.9%) |
| Secondary (8-15 years) | 31752<br>(54.8%) | 30514<br>(55.0%) | 62266<br>(54.9%) |
| Tertiary (16+ years) | 18326<br>(31.6%) | 17127<br>(30.9%) | 35453<br>(31.2%) |
| *Income Quintiles* |  |  |  |
| Poorest 20% | 7955<br>(13.7%) | 9123<br>(16.4%) | 17078<br>(15.1%) |
| Second 20% | 8952<br>(15.4%) | 10074<br>(18.2%) | 19026<br>(16.8%) |
| Middle 20% | 10676<br>(18.4%) | 11024<br>(19.9%) | 21700<br>(19.1%) |
| Fourth 20% | 12623<br>(21.8%) | 12123<br>(21.8%) | 24746<br>(21.8%) |
| Richest 20% | 17761<br>(30.6%) | 13149<br>(23.7%) | 30910<br>(27.2%) |
| *Employment status* |  |  |  |
| Unemployed | 12511<br>(21.6%) | 21141<br>(38.1%) | 33652<br>(29.7%) |
| Full time | 34125<br>(58.9%) | 21551<br>(38.8%) | 55676<br>(49.1%) |
| *Part time* | 11331<br>(19.5%) | 12801<br>(23.1%) | 24132<br>(21.3%) |
| *Use social media (frequency)* |  |  |  |
| *0: Never* | 10630<br>(18.3%) | 11089<br>(20.0%) | 21719<br>(19.1%) |
| *1* | 2562<br>(4.4%) | 2476<br>(4.5%) | 5038<br>(4.4%) |
| *2* | 3759<br>(6.5%) | 3585<br>(6.5%) | 7344<br>(6.5%) |
| *3* | 6449<br>(11.1%) | 6245<br>(11.3%) | 12694<br>(11.2%) |
| *4* | 21849<br>(37.7%) | 21070<br>(38.0%) | 42919<br>(37.8%) |
| *5* | 5565<br>(9.6%) | 5247<br>(9.5%) | 10812<br>(9.5%) |
| *6: several times an hour* | 7153<br>(12.3%) | 5781<br>(10.4%) | 12934<br>(11.4%) |
| *Global North or South* |  |  |  |
| *Global south* | 37732<br>(65.1%) | 34803<br>(62.7%) | 72535<br>(63.9%) |
| *Global north* | 20235<br>(34.9%) | 20690<br>(37.3%) | 40925<br>(36.1%) |
| *Country vulnerability* |  |  |  |
| *Resilient* | 40399<br>(69.7%) | 39978<br>(72.0%) | 80377<br>(70.8%) |
| *Vulnerable* | 17568<br>(30.3%) | 15515<br>(28.0%) | 33083<br>(29.2%) |

**Table 2.** Coefficient estimates of fixed effects model, random effects model and placebo test model.

|  | Fixed<br>effect | Random<br>effect | Placebo<br>fixed eff. |
| --- | --- | --- | --- |
| *Belief in*<br>*climate change* | 0.116***<br>(0.00) | 0.118***<br>(0.00) | -0.0126<br>(0.54) |
| *Age* | 0.0015***<br>(0.00) | 0.0015***<br>(0.00) | 0.0018***<br>(0.00) |
| *Age square* | -0.00003***<br>(0.00) | -0.00003***<br>(0.00) | -0.00003***<br>(0.00) |
| *Sex, Female* | 0.0249***<br>(0.00) | 0.0249***<br>(0.00) | 0.0259***<br>(0.00) |
| *Education* |  |  |  |
| *Secondary* | -0.0401***<br>(0.00) | -0.0400***<br>(0.00) | -0.0206***<br>(0.00) |
| *Tertiary* | -0.0806***<br>(0.00) | -0.0807***<br>(0.00) | -0.0512***<br>(0.00) |
| *Income quint.* |  |  |  |
| *Quintile 2* | -0.0260***<br>(0.00) | -0.0260***<br>(0.00) | -0.0256***<br>(0.00) |
| *Quintile 3* | -0.0458***<br>(0.00) | -0.0458***<br>(0.00) | -0.0443***<br>(0.00) |
| *Quintile 4* | -0.0563***<br>(0.00) | -0.0564***<br>(0.00) | -0.0529***<br>(0.00) |
| *Top quintile* | -0.0646***<br>(0.00) | -0.0647***<br>(0.00) | -0.0602***<br>(0.00) |
| *Employment* |  |  |  |
| *Full time* | 0.00420<br>(0.18) | 0.00418<br>(0.19) | 0.00472<br>(0.13) |
| *Part time* | 0.0560***<br>(0.00) | 0.0563***<br>(0.00) | 0.0578***<br>(0.00) |
| *Social media* | 0.0047***<br>(0.00) | 0.0046***<br>(0.00) | 0.0075***<br>(0.00) |
| *Covid case log* |  | 0.00913*<br>(0.02) |  |
| *North* |  | -0.0253<br>(0.26) |  |
| *Income, log* |  | -0.0101<br>(0.42) |  |
| *Constant* |  | 0.196<br>(0.06) |  |
| *N* | 113460 | 113460 | 113460 |

*p*-values in parentheses
\* $p < 0.05$ , \*\* $p < 0.01$ , \*\*\* $p < 0.001$

**Table 3.**
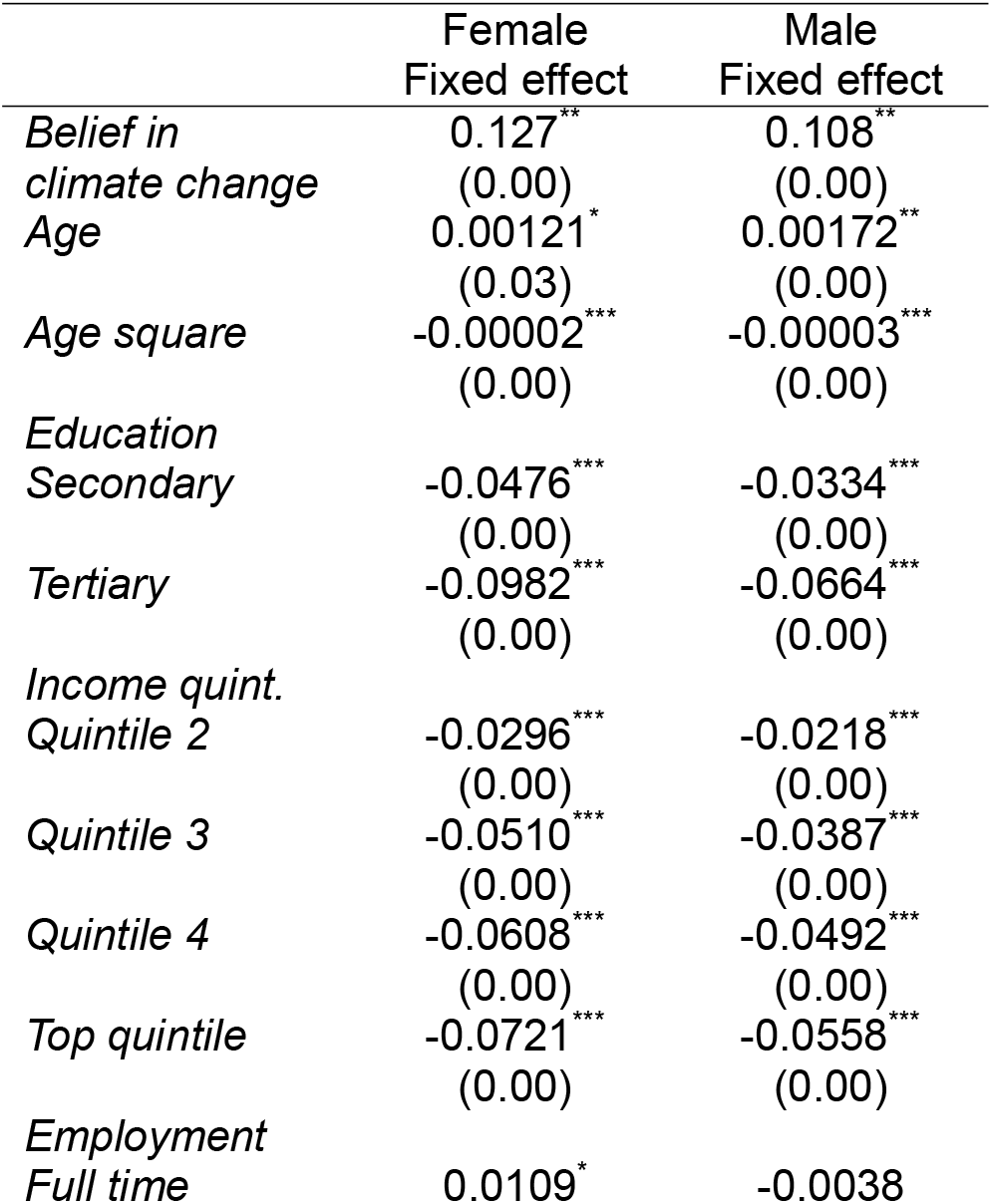
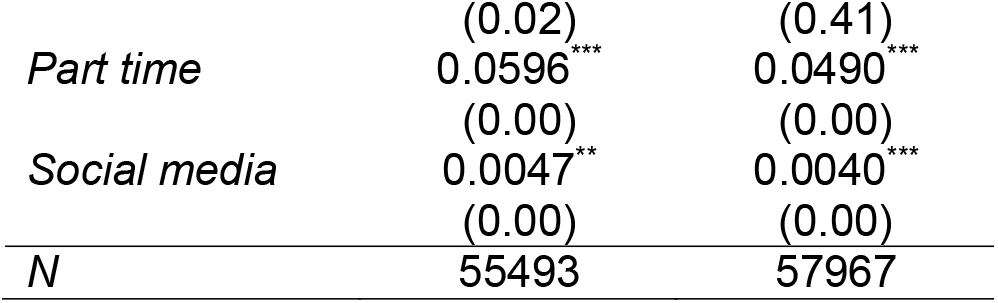
Heterogeneity across gender: instrumental variable fixed effects model.

The sample is made up of mostly males (51%) with reported extreme anxiety at 19% (males) and 22% (females). Age ranges from 15 to 99 with mean of nearly 40 years. Today most people belief in climate change as a major risk (56%) and nearly one in five never used social media in the past month; see table 1.

The fixed effect model shows that personal belief in climate change significantly raises the probability of extreme anxiety by 12% (95% confidence interval 6 – 17%). The test of endogeneity of personal belief rejects it being exogenous (the null hypothesis) with χ^2^ = 10.581 and p = 0.001. This also suggests that the coefficients are consistent and ignoring the endogeneity would have yielded inconsistent coefficients even though these are more efficient. Nevertheless, comparing the magnitudes of the two sets of coefficients side by side suggests that the differences are slight (mostly on the third digits). In such cases the differences make no practical or clinical difference.

The test of relevance of instruments (F = 115; p <0.0001) and the test of weak instruments is comfortably beyond the Stock-Yogo threshold of 19.9 (Cragg-Donald Wald F statistic = 526.3). Moreover, the overidentification test suggests restraint – both instruments i.e. beliefs in science two years ago and today are needed (Sargan-Hansen statistic = 0.367 and p = 0.54). In sum, all these tests point to a causal effect of belief in climate change on extreme anxiety. A local average treatment effect interpretation suggests that this effect is experienced by those who would be moved to belief in climate change due to an increase in the country average of belief in science.^27^

The other covariates can be interpreted briefly as they are in accord with recent empirical studies of social determinants of common mental disorders in rich and low income countries examined using standard depression measure.^19–21,23^ There is a non-linear effect of age and females show a higher likelihood of extreme anxiety, with a positive coefficient (confirmed by the heterogeneity result to follow). Higher educational attainment reduces the likelihood of extreme anxiety. Tertiary education shows a stronger protective effect (around 8%) compared to primary education or less. Higher income groups are less likely to report extreme anxiety, with the effect becoming more pronounced in higher quintiles. For example, the wealthiest fifth have coefficients around -6%. Those in part-time employment report significantly more extreme anxiety, compared to full-time employment or not in employment. Frequent social media use is significantly correlated with anxiety though the magnitude is small (0.5%).

It is well known that instrumental variable analysis relies on the untestable assumption of exogeneity of instruments and the literature has suggested placebo tests as evidence that would support such an assumption.^27,28^ If the placebo implied by the assumption indeed has null effect then confidence in the exogeneity assumption is enhanced. The treatment here is personal belief in climate change – if this were replaced by a placebo of a random person’s belief (a person drawn from the same country) then the placebo’s effect should be null. That is indeed the case (last column: -0.01%; p = 0.54).

The examination of heterogeneity is done across the sexes. The pattern of personal belief in climate change being significant and deleterious for mental health is repeated among men and women separately. There is no qualitative heterogeneity between them.

### Mechanism results

To help interpretation of this model and the mechanism I plot predicted probabilities of anxiety against the context of high or low vulnerability (vulnerable or resilient) to global warming, distinguishing those who believe in global warming as a major risk from those who do not. The plot in figure 5 draws predicted probabilities of experiencing extreme anxiety based on levels of vulnerability to climate change and personal beliefs about its risks. The vertical axis represents the probability of reporting extreme anxiety, while the horizontal axis identifies individuals as living in either in vulnerable or resilient contexts. Two lines represent believers and doubters of climate change.

**Figure 5.**
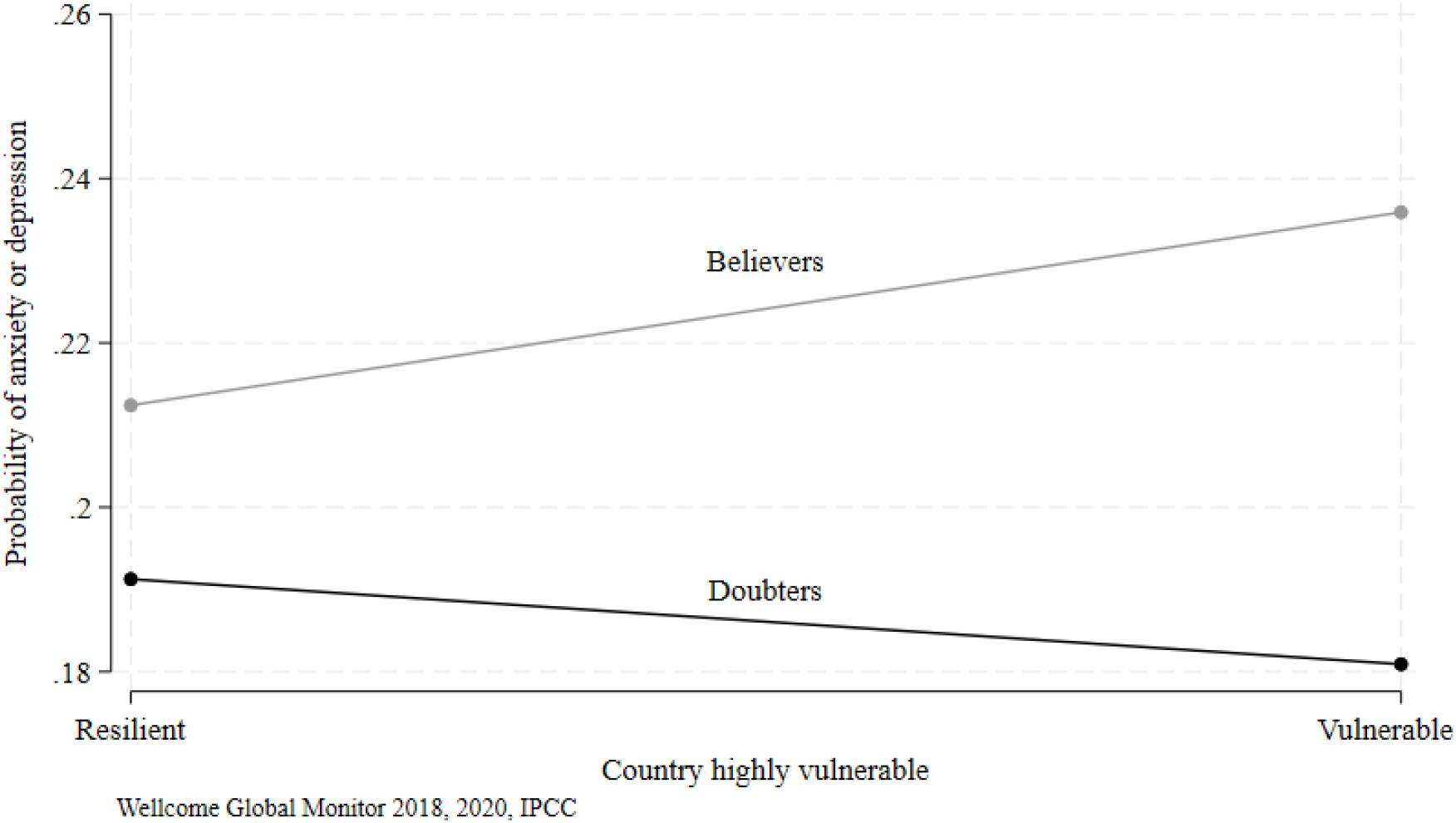
Marginal plot of probabilities of extreme anxiety by belief in climate change, in vulnerable and resilient countries. Source: Wellcome Global Monitor (2023).

The key insight from this plot aligns with the model interpretation above: belief in the serious risk posed by global warming is strongly associated with an increased probability of extreme anxiety. For individuals who view global warming as a major threat, the probability of extreme anxiety is consistently higher across both vulnerable and resilient contexts. This group shows a pronounced increase in anxiety and depression as vulnerability increases, with the probability rising from approximately 21% in low-vulnerability contexts to over 24% in high-vulnerability contexts. This trend suggests that those who already view global warming as a serious issue are particularly susceptible to anxiety when facing high environmental vulnerability. This insight is obtained by modelling personal and contextual factors which has discovered a more accentuated difference only hinted in the plot of country averages in figure 4.

In contrast, individuals who do not perceive global warming as a major risk exhibit a stable probability of experiencing extreme anxiety, irrespective of where they live, either in vulnerable or resilient nations. This lower and relatively stable probability across varying levels of vulnerability indicates that for doubters of climate change anxiety and depression are less influenced by environmental context.

This interaction effect reinforces the model’s finding that belief in climate risk amplifies the mental health impact of high vulnerability contexts. The psychological burden of climate change appears most acute among those who are both aware of its risks and live in highly vulnerable environments, potentially due to perceived personal or societal risk. Those without this belief appear less affected, suggesting a buffering effect against climate-related anxiety.

Overall, this plot emphasizes how the interaction between personal beliefs about climate change and contextual vulnerability can shape mental health outcomes. While vulnerability alone does not increase extreme anxiety for those who doubt climate risks, it significantly impacts those who acknowledge the threat, underscoring the importance of targeted support in vulnerable populations. This is a new finding that deserves discussion.

## DISCUSSION

Personal belief in climate change as a major risk is found to have an effect on extreme anxiety, sufficient to hinder normal daily activities for a fortnight. This is among the first pieces of evidence on general anxiety arising from a *specific* belief in climate change. The finding has important implications. Recall that the physics of climate change implies that climate change is not going away soon (say this decade) because of the lag between the accumulated *stock* of greenhouse gases and their effects on global temperatures, rainfall patterns and sea level heights – it is primarily the stock and not the current flow of greenhouse gases that matters in secular time.^2^ With these secular changes in the earth temperatures, rainfall patterns, and sea level heights (their frequency and intensity ramped up) more people will come to believe in climate change as a major risk. Therefore the evidence suggests more widespread anxiety in the world population will follow.

The analysis also furnishes evidence to elucidate one mechanism behind the cause and effect in terms of dissonance with what actions are collectively called for by the Panel. It is the dissonance that establishes the link and when the dissonance is removed or, conversely, when nation resilience is increased, the link also weakens. This has a bearing on public policy.^29^

### Counsels of despair

A sense of despair at times creeps in when one reads debates on climate change and climate actions with a view towards public policy.^29^ The tipping point to bend the curve of climate change is deemed to have passed by some, tempting protagonists to abandon hope.^30,31^ This analysis does not impinge on this counsel but there is an interpretation of these new findings that may prompt another counsel of despair. The analysis shows that believers in climate change have higher probabilities of experiencing extreme anxiety. Its converse may imply that in order to preserve mental health one may choose to doubt climate change. But this would be a second counsel of despair. A better implication is furnished by the mechanistic analysis. It suggests that resilient nations, compared to vulnerable nations, are home to residents who suffer *less* from extreme anxiety even with the same belief in climate change. This is the more hopeful message from the data. Instead of abandoning belief in climate change, collective actions that are effective by governments, institutions and persons are called for. Some may hold a basis for the first counsel of despair, “abandon all hope for we have passed the tipping point”.^30^ But there is even less basis for the second counsel because the analysis strongly suggests that effective climate actions that build resilience across nations can evidently yield mental health benefits today in reduced extreme anxiety.

### Sustainable development goals synergy

As the world approaches the finish line of sustainable development goals, interlinkages between the seventeen goals take centre stage along with their contradictions and synergies.^17,31^ The authors systematically reviewed the empirical literature to tease out instances of goals that are conflicting and those that are mutually supporting. Given the complexity of global development challenges and the breadth of the set of goals, it is clear that empirics will have a strong voice on the evaluation of achievements of the goals. These new results discover an empirical synergy between the goals of health for all and urgent actions on climate change. The mechanism posited above captures the logic of the synergy, when urgent actions on climate change are taken to reduce national vulnerability then personal belief in climate change weakens as a cause that drives extreme anxiety. Actions in pursuit of one goal yield welcome progress on two fronts: health and climate.

No doubt encouraged by these goals, the recent literature on climate anxiety has been growing across the world.^9–11^ The new results complement this literature in three ways. First, it resonates with the deleterious effect on anxiety brought about by climate change while at the same time broadening the literature by using a general (non-specific) anxiety question. Second, it broadens the evidence base to the largest number of countries (108) for this kind of investigation. Last, it adds to a new driver of anxiety, specifically personal belief in climate change.

The data point to some **limitations** as hinted above. First, believers in climate change are apt to act and thereby reduce the dissonance between collective actions and perceived required actions, and this may be self-efficacious which in turn may bring about reduction in extreme anxiety.^32^ Unfortunately, the Wellcome Trust did not collect this information.^18^ Apart from this lack of information on climate action, consumption of disinformation or misinformation, say through social media, is also lacking. Tobacco smoking followed by climate change are two sources of health harms which have been subject to vigorous public and political debate on dis/misinformation.^3,4,16^ Again, no information is collected on consumption dis/misinformation such as hurricane Milton being geoengineered by the U.S. government (<u>The Independent</u> 9 October 2024; <u>NBC</u> News 10 October 2024). Had information about action and dis/misinformation been available the analysis would have been sharper in attributing the effect (extreme anxiety) to personal belief in climate change. Last, the outcome is coarse as elicited with only one question but also because the *number* of days lost to extreme anxiety was not asked, only whether there had been any of those periods. Even so, a binary question was proven to be cost-effective in showing the harmful effect of personal belief in climate change. And importantly, it is determined by similar factors often found significant when a standard measure of depression is used, a form of external validation (see the brief interpretation above). Using a more complex climate anxiety scale (up to 22 questions) may be preferable as it would accumulate findings on a climate anxiety scale. Tangentially, an equally general scale of depression (not specifically primed with climate change issues) could be used, in particular the Center [sic] of Epidemiologic Scale – Depression which has been widely used in rich and low-income countries.^20,21^

The **strengths** are numerous, not least the world coverage and the person sample size. This has lent power to the analysis which yielded strong inference on the effect and associations. This also has enabled generalisations to be made across a wide range of country characteristics that are time invariant. For example, irrespective of a country’s economic development in 2020 (expressed in the fixed effects), the effect of personal belief on the probability of extreme anxiety is obtained above. Or regardless of the country location on the planet, the effect of personal belief is the same significant magnitude as above. The coverage of age range is also unusually wide, from teenagers to older adults up to 99. This allows demographic generalisation. There may be variations across age (recall Greta Thunberg) but even then, it is still true that personal belief in climate change as a major risk leads to an increase in the probability of extreme anxiety. Last, the analysis attempted to fortify the associations by using a more robust method of instrumental variable analysis, and complementing it with a placebo test and a mechanism analysis. The work goes beyond estimating associations, though that is also done. Due to the lack of information that may be common (see weaknesses above), applying the more robust method is critical.

Given the global nature of climate change which goes hand in hand with the complexity of the risks it poses this work reminds us to reaffirm the importance of science and the belief in science. The inter-related and long-term changes inherent to climate change can only be effectively understood and responded to with science as our ally. It turns out that even in the short term, accepting such a quintessentially scientific statement on climate change can have a personal health effect. The mechanism at work behind this effect offers another hope: it reinforces the motivation to take urgent collective actions on climate change.

## Data Availability

All data used are available from the Wellcome Trust website.

https://ipcc-browser.ipcc-data.org/browser/dataset?id=3736

## Notes

### Competing Interest Statement

The authors have declared no competing interest.

### Funding Statement

This study did not receive any funding

### Author Declarations

University of Manchester Ethics Committee waived ethical approval for this work in its use of publicly available deanonymised data.

